# Systematic mediation and interaction analyses of kidney function genetic loci in a general population study

**DOI:** 10.1101/2023.04.15.23288540

**Authors:** Dariush Ghasemi-Semeskandeh, David Emmert, Eva König, Luisa Foco, Martin Gögele, Laura Barin, Ryosuke Fujii, Christian Fuchsberger, Dorien J.M. Peters, Peter P. Pramstaller, Cristian Pattaro

## Abstract

Chronic kidney disease (CKD) is a complex disease affecting >10% of the global population, with large between-and within-continent variability reflecting major environmental effects. To identify molecular targets for treatment, genome-wide association study meta-analyses (GWAMAs) of CKD-defining traits have identified hundreds of genetic loci in aggregated populations. However, while GWAMAs estimate the average allelic effect across studies, single population studies may be relevant to unravel specific mechanisms. To assess whether an individual study from a specific population could extend existing knowledge on kidney function genetics, we selected 147 kidney function relevant loci identified by a large GWAMA, assessing their association with the glomerular filtration rate estimated from serum creatinine (eGFRcrea) in the Cooperative Health Research In South Tyrol (CHRIS) study (n=10,146), conducted in an Alpine region where thyroid dysfunction is common. We replicated associations with single nucleotide polymorphisms (SNPs) at 11 loci, showing up-to-5.4 times larger effect sizes than in the corresponding GWAMA, not explainable by minor allele frequency differences. Systematic mediation analysis across 70 quantitative traits identified serum magnesium and the activated partial thromboplastin time as partial mediators of the eGFRcrea associations at *SHROOM3* and *SLC34A1*, respectively. Given that free triiodothyronine and thyroxine were effect modifiers across all loci, we conducted SNP-by-thyroid stimulating hormone (TSH) interaction analyses, identifying significant interactions at *STC1*: SNPs had larger effects on eGFRcrea at higher TSH levels, possibly reflecting stanniocalcin-1 autocrine and paracrine role. Individual population studies can help characterize genetic associations. The interplay between phenotypes at *SHROOM3* and *SLC34A1* and the role of thyroid function as a genetic effect modifier warrant further investigations.

## Introduction

Chronic kidney disease (CKD) is a common complex disease that increases the risk of kidney failure, cardiovascular mortality, and all-cause mortality^1^. CKD is predicted to become the 5^th^ leading cause of death by 2040^2^. While affecting >10% of the population globally, CKD prevalence shows marked variability across countries, both between and within continents^3^. Between-continent variability is likely guided by combinations of environmental and genetic differences, such as is the case with sickle cell trait and albuminuria^4^. Within-continent variability could be more related to environmental differences, including lifestyle, public health policies, and assessment methods^4,5^.

To unravel the genetic basis of CKD, several genome-wide association study meta-analyses (GWAMAs) were conducted that analyzed the CKD-defining marker glomerular filtration rate (GFR) estimated from serum creatinine (eGFRcrea)^6^. More than 400 associated loci have been identified to date^7^. While GWAMAs estimate the average allelic effect across studies, evidence shows that individual population studies may be extremely relevant to advance discoveries not only in the field of rare diseases^8^ but also with regard to markers of common chronic diseases such as triglycerides and low density lipoprotein (LDL) cholesterol^9^, age at diabetes onset^10^, and mood disorders^11^. The evidence accumulated so far suggests that there is no general rule and that the contribution of an individual study to the dissection the biological basis of common phenotypes can only be assessed on a case-by-case basis and depends on study-specific characteristics.

Here we analyzed genetic associations with eGFRcrea in the Cooperative Health Research In South Tyrol (CHRIS) study, a general population study conducted in a mountainous region of central Europe^12^. A sample from this same population was previously instrumental to identify genetic linkage of serum creatinine with the *APOL1*-*MYH9* locus^13^ and to replicate common variants associated with eGFRcrea^14^. Characteristics of the geographical region where the study was conducted include the demographic stability over time^15^, the rural environment, and a high frequency of hypothyroidism, typical of mountainous regions^16^. Aim of our analysis was to assess whether this particular study sample could provide information that extend current knowledge on kidney function genetics, by leveraging on local specific characteristics.

For a given single population study, with a relatively limited sample size, to help expand knowledge on loci already identified by a large consortium, it is necessary and appropriate to limit investigation to the subset of loci associated with the phenotype of interest in the given study. Thus, we first identified which, among 147 loci found to be associated with kidney function in a large GWAMA from the CKDGen Consortium^17^, were associated with eGFRcrea in the CHRIS study. We take it for granted that the identified loci are relevant for kidney function. This was demonstrated by Wuttke et al.^17^ through the combined analysis of eGFRcrea and blood urea nitrogen (BUN).

CKDGen loci were tested for replication in the CHRIS study. The purpose of this replication analysis was to identify those loci for which a study of limited size can try to contribute new information. The replication analysis should not be understood as a demonstration of the validity of the analyzed loci: in fact, they should already be considered as validated, as they were identified through a large meta-analysis on more than 760,000 individuals and replicated in an independent study on more than 280,000 individuals^17^.

The features of the replicated loci were then assessed against corresponding European ancestry CKDGen GWAMA results. Given that several replicated loci showed larger effects in CHRIS than in the European ancestry CKDGen GWAMA, we examined the possible reasons of such larger effects and conducted extensive mediation analyses across 70 quantitative biochemical and anthropometric traits, reflecting multiple health conditions. Further, after observing a systematic modification of the genetic association with eGFRcrea when adjusting for thyroid-related traits, we conducted SNP-by-thyroid function interaction analyses. The aims of the mediation and the interaction analyses were to identify the presence of intermediate phenotypes and effect modifiers, respectively, that were specific of the study sample.

## Methods

### Study sample

The present analyses were based on 10,146 individuals with complete genotype data who participated to the CHRIS study, a population-based study conducted in South Tyrol, Italy, between 2011 and 2018, which has been extensively described elsewhere^12,18^. Briefly, participants underwent blood drawing, urine collection, anthropometric measurements, and clinical assessments early in the morning, after an overnight fast. Medical history was reconstructed through interviewer-and self-administered standardized questionnaires. Drug treatment was identified via barcode scan of the drug containers that participants were instructed to bring at the study center visit, with the drug code linked to an official drug databank^12^.

### eGFRcrea estimation

Serum creatinine (SCr) was measured with a colorimetric assay on Roche Modular PPE (n=4,176) and Abbott Diagnostic Architect c16000 (n=5,970) instrumentations. According to previous analyses^18^, SCr was normalized by instrument (fixed effect) and participation period (random effect) using a linear mixed model implemented in the ‘lme4’ R package^19^. To reflect the same methods used in the CKDGen Consortium meta-analysis^17^, eGFRcrea was estimated using the 2009 CKD-EPI equation^20^ implemented in the R package ‘nephro’ v1.2^21^ and winsorized at 15 and 200 ml/min/1.73m^2^. Finally, the natural logarithm transformation was applied.

### Genotype imputation

Genotyping was performed in two batches using genotyping array chips based on the Illumina Human Omni Microarray platform. Following quality control analysis, the two data batches were merged, phased with SHAPEIT2 v2.r837, using the duoHMM method (--duohmm -W 5) with 800 states and 30 rounds^22^, and imputed based on the TOPMed r2 standard reference panel on the Michigan Imputation Server^23^. We obtained 34,084,280 SNPs with minimum imputation quality index Rsq ≥0.3 and minor allele count (MAC) ≥1, aligned with human genome assembly GRCh38. A genetic relatedness matrix (GRM) was obtained from the autosomal genotyped SNPs using EPACTS v3.2.6 modified for compatibility with assembly GRCh38. Genetic principal components (PCs) were estimated based on the genotyped variants data with minor allele frequency (MAF) >0.05 using GCTA^24^.

### GWAS and replication of CKDGen results

We conducted a GWAS of age-and sex-adjusted residuals of ln(eGFRcrea) using the EMMAX method^25^ implemented in EPACTS v3.2.6, including the GRM to model the sample structure. After the analysis, we removed variants with minor allele frequency (MAF) <0.005, leaving 10,158,100 SNPs for further characterization. Genomic inflation was assessed on this set of SNPs by estimating the genomic control factor λ^26^.

Summary statistics from the CKDGen GWAMA^17^ were downloaded from the publicly available repository at https://ckdgen.imbi.uni-freiburg.de. Given a subset of 4661 samples from the CHRIS study was previously included in the CKDGen GWAMA, we removed CHRIS data from the CKDGen GWAMA summary statistics using MetaSubtract v1.60^27^. After this process, the 147 variants remained genome-wide significant in CKDGen. We lifted the CKDGen genomic positions from the GRCh37 to the GRCh38 map, using CrossMap v0.5.3^28^.

Given small-sample bias can cause large effect estimate fluctuations and given it is not uncommon to observe small studies with opposite effect directions compared to the meta-analysis results in large GWAMAs, to ensure consistency of the effect direction, replication of the 147 loci in CHRIS was assessed based on a one-sided test, following common practice^29^. Statistical significance for replication was set at the Bonferroni-corrected level of 0.00034 (0.05/147), for any variant in strong linkage disequilibrium (LD) (r^2^>0.8) with a lead CKDGen variant. LD was estimated using emeraLD v0.1^30^. The total number of SNPs within the 147 loci was 6337. Adjustment for 147 independent tests covers 98% of the SNP’s cumulative variance (Suppl. Fig. S1).

We compared the variance of ln(enforcers) explained by SNPs in CHRIS versus CKDGen European Ancestry analysis as b^2^ × 2 × p × (1-p) / var(y),^17^ where y is the age-and sex-adjusted ln(eGFRcrea), b is the estimated SNP effect on y, and p is the SNP allele frequency. For comparison with CKDGen, we used a common estimate of var(y).

### Mediation analysis

Replicated ln(eGFRcrea)-SNP associations were submitted to mediation analysis across 70 quantitative anthropometric, blood pressure and biochemical traits (listed in Supplementary Table S1), following the flowchart represented in Fig. 1. SCr was included as a positive control. To prevent potential measurement instrument effects^18^, we applied quantile normalization to each trait (Supplementary Methods). Mediation analysis was conducted for each *trait* following the established 4-step framework and fitting linear regression models throughout:

**Figure 1.**
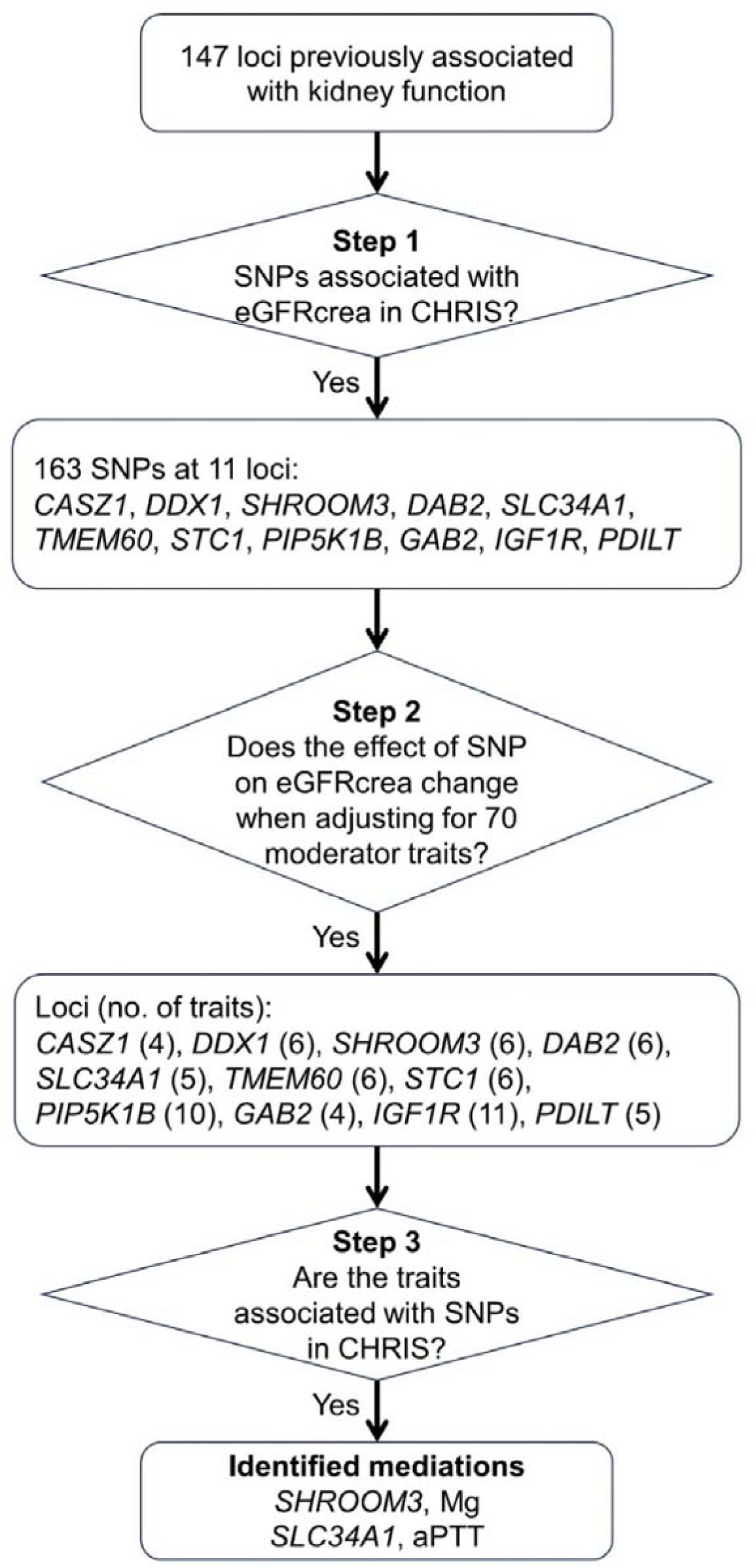
Flowchart of the mediation analysis. Abbreviations: Mg, serum magnesium; aPTT, activated partial thromboplastin time.

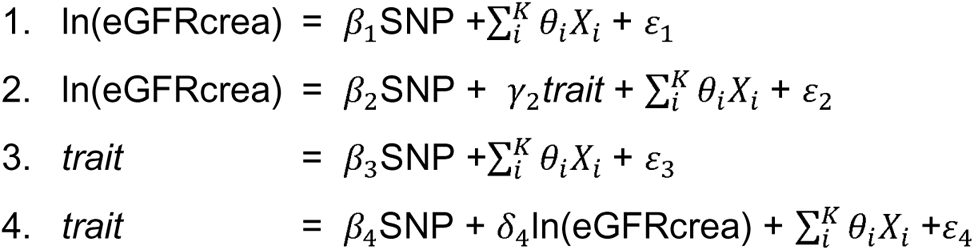

where ε_1_ to ε_4_ refer to Gaussian error terms, and ∑_*i*_^*K*^ *θ_i_* X*_i_* zindicate the inclusion of K covariates, namely age, sex, the first 10 genetic PCs and an intercept. Fitting PC-adjusted simple linear models came for practical reasons after observing substantial equivalence with kinship-adjusted linear mixed models obtained with EMMAX (Suppl. Fig. S2). Step 1 corresponds to the GWAS results: given we investigated the replicated variants, the estimate of β_1_ (b_1_) was always significant. We defined two criteria that must be satisfied in order for a trait to be considered at least a partial mediator of the ln(eGFRcrea)-SNP relation: (i) evidence of a substantial alteration of the SNP effect on ln(eGFRcrea) when adjusting for the trait (step 2) and (ii) the SNP is also associated with the trait (step 3) at a multiple-testing corrected level of 6.5×10^-5^ (corresponding to 0.05 / (70 traits ×11 independent loci)). Criterion (i) was assessed by analyzing the distribution of the estimate of β_2_ (b_2_): for a specific trait, b_2_ was classified as an outlier according to the rule b_2_ < P_10_ – 1.5 (P_75_ – P_25_) or b_2_ > P_90_ + 1.5 (P_75_ – P_25_), where P_10_, P_25_, P_75_, and P_90_ indicate the 10^th^, 25^th^, 75^th^, and 90^th^ percentile of the distribution of b_2_ across all traits. This is essentially a more stringent version of the Tukey’s rule for outlier detection.

To support associations in step 3 and assess whether lack of mediation evidence might have been due to lack of power in the CHRIS study, we interrogated the SNP-trait associations that passed the mediation analysis step 2 on the PhenoScanner v2^31^ (http://www.phenoscanner.medschl.cam.ac.uk/, interrogated on 31-Jan-2023), GWAS Catalog^32^ (https://www.ebi.ac.uk/gwas/docs/api; 24-Feb-2023), and the ThyroidOmics Consortium summary statistics^33^ (https://www.thyroidomics.com), for associations at *P*<5.0×10^-8^.

### Interaction with thyroid function phenotypes

Free triiodothyronine (FT3) and thyroxine (FT4) did not qualify as mediators but caused a major departure of b_2_ from its expected distribution (see Results). These traits may reflect thyroid gland issues. Given the relevance of hypothyroidism in the region, we analyzed these traits in detail. Because FT3 and FT4 levels were measured only when the thyroid stimulating hormone (TSH) level was <0.4 or >3.8 µUI/mL, we started with a series of sensitivity analyses to exclude the presence of underlying sample stratification. First, we tested whether the distribution of the genotypes was different between individuals with and without measured FT3 and FT4, using a two-sided Wilcoxon test at a significance level of 0.05. Second, to verify whether there was any geographical cluster of individuals with extreme TSH levels (and so measured FT3 and FT4 levels), which might have implicated population stratification, we further adjusted the ln(eGFRcrea)-SNP association model in step 1 for municipality of residence.

After excluding the presence of sample stratification, we tested the interaction between the SNPs and TSH levels, which were measured in the whole sample. We excluded 416 individuals reporting any condition among thyroid cancer (n=16), kidney cancer (n=1), goiter (n=277), having undergone surgery to the thyroid gland (n=312), or having both missing TSH measurement and therapy information (n=4), leaving 9730 individuals for the interaction analysis. Regression models included both the main and the interaction effect terms and were adjusted for the same covariates used in the step 1 model. In addition to quantitative TSH levels, we also tested interaction with broadly defined hyper-and hypothyroidism. Hyperthyroidism was defined as a TSH level of <0.4 µUI/mL or use of thiamazole (n=3) or propylthiouracil (n=1). Hypothyroidism was defined as TSH levels >3.8 µUI/mL or reported use of levothyroxine sodium (n=512). No other thyroid-related treatment was reported. The statistical significance level for interaction testing was set at 0.0045=0.05/11 loci.

### Ethical statement

The CHRIS study was approved by the Ethical Committee of the Healthcare System of the Autonomous Province of Bozen/Bolzano, protocol no. 21/2011 (19 Apr 2011). All participants provided written informed consent. All the methods were performed in strict accordance with the approved protocol.

## Results

In the study sample, the median age was 46.5 years (interquartile range, IQR: 32.8, 57.6), females were 55%, and the median eGFRcrea level was 92.2 (IQR: 81.3, 103.2) ml/min/1.73m^2^ (Table 1; additional clinical characteristics are reported in Supplementary Table S1). The genomic inflation factor λ was 1.02 (Supplementary Fig. S3), supporting appropriate modeling of the sample structure. In the CHRIS study, we replicated 11 of the 147 CKDGen loci (1-sided P-value between 3.34×10^-4^ and 4.51×10^-7^; Table 2; Supplementary Table S2). For each replicated locus we retained all variants in LD (r^2^>0.8) with the lead SNP, totaling 163 variants over the 11 loci (Supplementary Table S3). The replicated loci displayed very similar LD pattern in CHRIS as in CKDGen (Supplementary Fig. S4). Among the replicated loci, variants’ effect magnitude at *CASZ1*, *DDX1*, *PIP5K1B*, *GAB2*, and *IGF1R* was significantly larger in CHRIS than in CKDGen (Fig. 2; Table 2). Replicated loci explained a larger proportion of ln(eGFRcrea) variance (Fig. 3A). For them, the CHRIS-to-CKDGen effect ratio varied between 1 and 5.4 and was not entirely explained by MAF differences (Fig. 3B; Supplementary Table S3). Three broad groupings characterized by MAF and effect size differences could be distinguished:

1. at *PDILT*, *SLC34A1* and *SHROOM3*, we observed similar effect magnitude, regardless whether MAF was similar or lower in CHRIS, consistent with the fact that these loci were amongst the ones identified by the earliest GWAS thanks to their large effects, counterbalancing the still relatively limited sample sizes and the coarser genomic imputation^34,35^;
2. at *DAB2*, *TMEM60*, *PIP5K1B*, *GAB2*, and *IGF1R*, MAF was similar or slightly larger in CHRIS (effects in CHRIS were 1.3-to-5 times larger than in CKDGen);
3. at *STC1*, *DDX1*, and *CASZ1*, we observed lower MAF in CHRIS and about 1.8-to-5.4 times larger effects in CHRIS.

**Figure 2.**
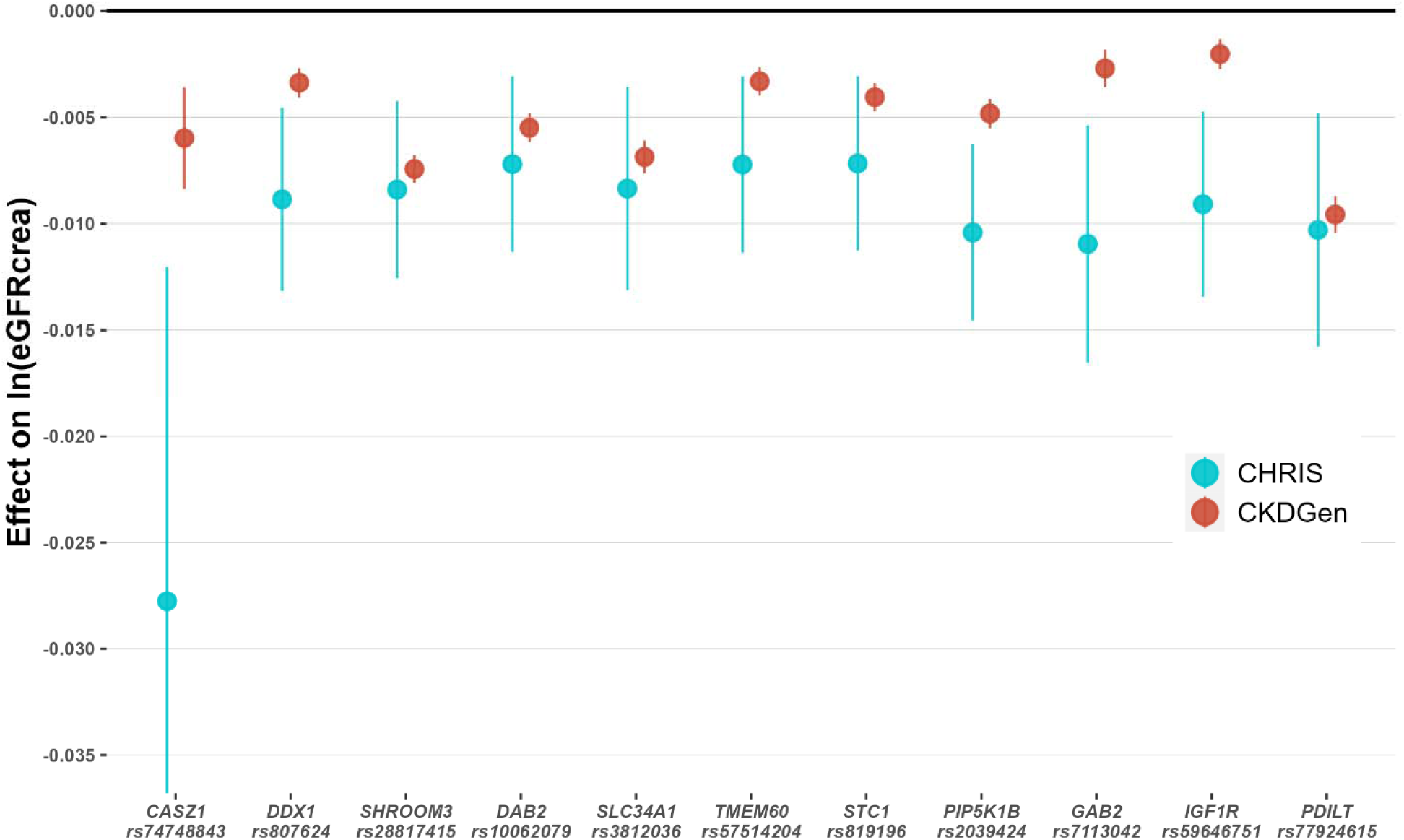
Effect sizes with 95% confidence intervals for the most associated variants at each locus in CKDGen and CHRIS following harmonization to the eGFRcrea-lowering allele. The effect is meant per copy of the effect allele on the natural logarithm of eGFRcrea in ml/min/1.73m^2^ (y-axis). For *CASZ1*, *DDX1*, *PIP5K1B*, *GAB2*, and *IGF1R* the magnitude of the effect in CHRIS was larger than in CKDGen.

**Figure 3.**
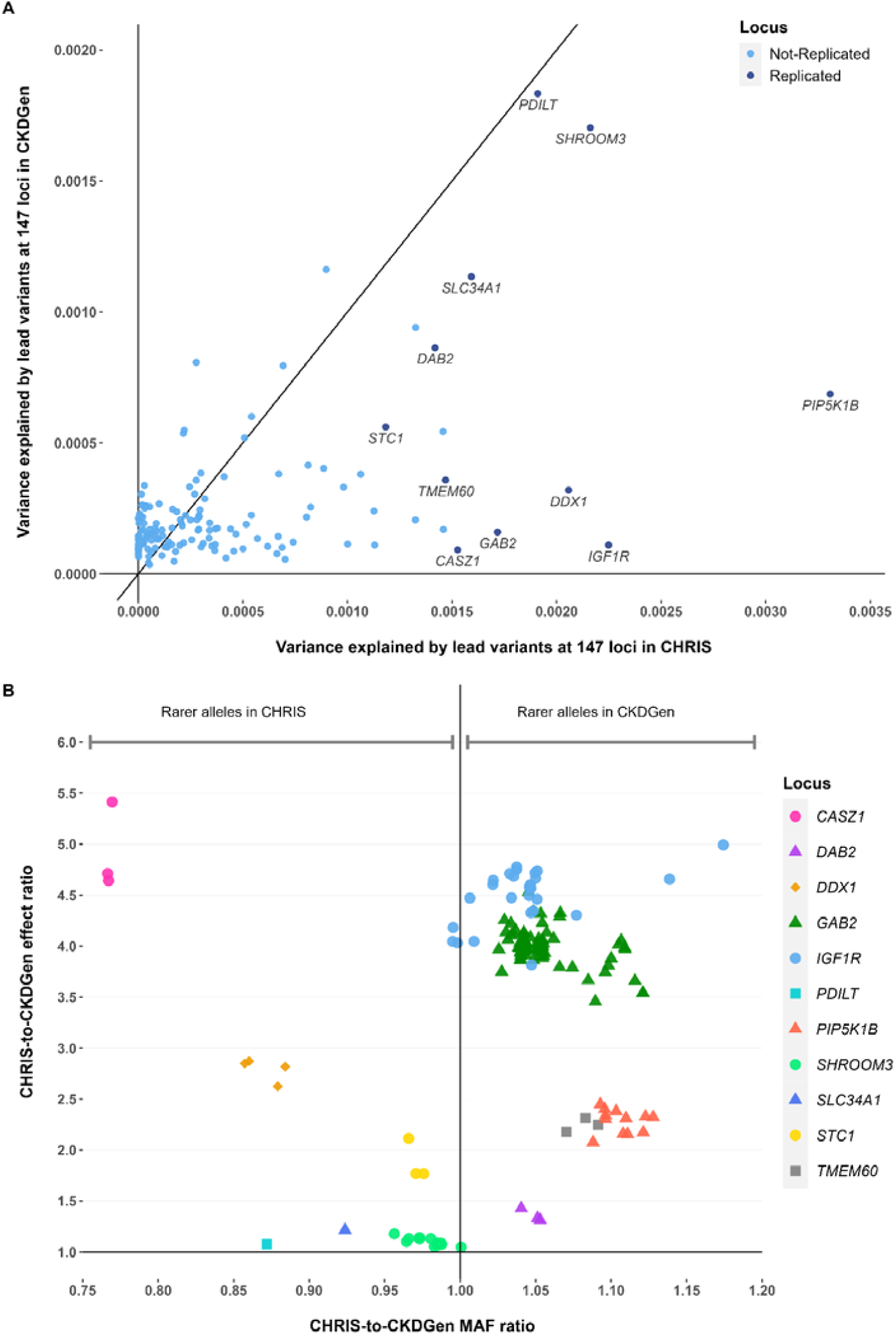
Comparison of the characteristics of all 163 variants in the 11 replicated loci in CKDGen versus CHRIS after alignment of the effect alleles. **Panel A**: Variance of age-and sex-adjusted ln(eGFRcrea) in CHRIS versus CKDGen EUR explained by the lead variants at each of the 147 loci. Replicated loci are labeled with the locus name. **Panel B**: The ratio between CHRIS and CKDGen minor allele frequencies (MAF, x-axis) is plotted against the ratio between CHRIS and CKDGen effect estimates (y-axis). Effects are expressed in terms of change of the natural logarithm of eGFRcrea in ml/min/1.73m^2^ per copy of the effect allele.

**Table 1.** Main characteristics of the 10,146 study individuals. Additional clinical characteristics are described in Supplementary Table 1.

| Participants' characteristics | Median (IQR) or N (%) |
| --- | --- |
| Age, years | 46.5 (32.8 – 57.6) |
| Females | 5,585 (55.1%) |
| eGFR<sub>crea</sub>, ml/min/1.73 m<sup>2</sup> (n=10,141) | 92.2 (81.3 – 103.2) |
| Mg, mg/dl (n=10,143) | 2.0(1.9 – 2.1) |
| aPTT, seconds (n=6,369) | 29.9 (28.6 – 31.4) |
| TSH, μUI/mL (n=10,142) | 1.38 (0.98 – 1.93) |
| FT3, pg/ml (n=466) | 3.00 (2.70 – 3.30) |
| FT4, ng/dl (n=469) | 0.98 (0.87 – 1.08) |
| Hyperthyroidism (TSH < 0.4 or drug treatment) | 172 (1.7%) |
| Normal (0.4 < TSH < 3.8 and no drug treatment) | 8,972 (88.5%) |
| Hypothyroidism (TSH > 3.8 or drug treatment) | 998 (9.8%) |

Abbreviations: IQR, Interquartile range; TSH, Thyroid-stimulating hormone; FT3, Free triiodothyronine; FT4, Free thyroxine; Mg, serum magnesium; aPTT, activated partial thromboplastin time.

**Table 2.** The 11 loci identified from the CKDGen Consortium GWAS that showed significant effects in CHRIS. Loci were considered replicated if any variant in strong LD with the CKDGen lead variant was significantly associated with ln(eGFRcrea) in CHRIS. Consequently, for each locus, reported are either the results on the CKDGen lead variant or the results of the replicated variant in the locus when the lead variant did not replicate itself (see Methods).

| Locus<br>(gene name) | CKDGen<br>lead SNP | Chr:Pos<br>(build 38) | EA/OA | CKDGen Trans-Ethnic<sup>1</sup> |  |  | CKDGen EUR<sup>1</sup> |  |  | CHRIS |  |  |  |
| --- | --- | --- | --- | --- | --- | --- | --- | --- | --- | --- | --- | --- | --- |
|  |  |  |  | EAF | b(SE)* | P-value | EAF | b(SE)* | P-value | EAF | b(SE) | 1-sided P-value | 2-sided P-value |
| **CASZ1** | rs74748843 | 1:10,670,853 | T/C | 0.071 | -0.00480<br>(0.00078) | $7.84 \times 10^{-10}$ | 0.021 | -0.00598<br>(0.00123) | $1.09 \times 10^{-6}$ | 0.016 | -0.02776<br>(0.00806) | $2.85 \times 10^{-4}$ | $5.70 \times 10^{-4}$ |
| **DDX1** | rs807624 | 2:15,642,347 | G/T | 0.580 | -0.00320<br>(0.00029) | $8.28 \times 10^{-28}$ | 0.660 | -0.00338<br>(0.00035) | $1.70 \times 10^{-21}$ | 0.701 | -0.00886<br>(0.00221) | $3.00 \times 10^{-5}$ | $5.99 \times 10^{-5}$ |
| **SHROOM3** | rs28817415 | 4:76,480,299 | T/C | 0.400 | -0.00730<br>(0.00029) | $3.17 \times 10^{-137}$ | 0.440 | -0.00744<br>(0.00034) | $7.32 \times 10^{-109}$ | 0.428 | -0.00840<br>(0.00214) | $4.21 \times 10^{-5}$ | $8.42 \times 10^{-5}$ |
| **DAB2** | rs10062079* | 5:39,393,631 | A/G | 0.039 | -0.00480<br>(0.00029) | $2.07 \times 10^{-60}$ | 0.430 | -0.00549<br>(0.00035) | $6.19 \times 10^{-56}$ | 0.453 | -0.00721<br>(0.00211) | $3.24 \times 10^{-4}$ | $6.49 \times 10^{-4}$ |
| **SLC34A1** | rs3812036 | 5:177,386,403 | T/C | 0.260 | -0.00650<br>(0.00039) | $2.96 \times 10^{-62}$ | 0.260 | -0.00687<br>(0.00040) | $2.31 \times 10^{-67}$ | 0.240 | -0.00836<br>(0.00245) | $3.27 \times 10^{-4}$ | $6.55 \times 10^{-4}$ |
| **TMEM60** | rs57514204* | 7:77,714,744 | T/C | 0.380 | -0.00340<br>(0.00029) | $3.53 \times 10^{-31}$ | 0.410 | -0.00332<br>(0.00034) | $1.48 \times 10^{-22}$ | 0.439 | -0.00723<br>(0.00212) | $3.34 \times 10^{-4}$ | $6.67 \times 10^{-4}$ |
| **STC1** | rs819196* | 8:23,885,208 | T/A | 0.410 | -0.00410<br>(0.00029) | $1.47 \times 10^{-44}$ | 0.450 | -0.00406<br>(0.00034) | $1.43 \times 10^{-33}$ | 0.437 | -0.00718<br>(0.00210) | $3.21 \times 10^{-4}$ | $6.41 \times 10^{-4}$ |
| **PIP5K1B** | rs2039424 | 9:68,817,258 | G/A | 0.360 | -0.00440<br>(0.00029) | $4.75 \times 10^{-51}$ | 0.380 | -0.00483<br>(0.00035) | $1.12 \times 10^{-42}$ | 0.421 | -0.01042<br>(0.00212) | $4.51 \times 10^{-7}$ | $9.01 \times 10^{-7}$ |
| **GAB2** | rs7113042* | 11:78,324,786 | A/G | 0.740 | -0.00270<br>(0.00039) | $4.62 \times 10^{-12}$ | 0.830 | -0.00271<br>(0.00045) | $1.86 \times 10^{-9}$ | 0.823 | -0.01096<br>(0.00286) | $6.41 \times 10^{-5}$ | $1.28 \times 10^{-4}$ |
| **IGF1R** | rs59646751 | 15:98,733,292 | T/G | 0.300 | -0.00230<br>(0.00029) | $3.97 \times 10^{-15}$ | 0.310 | -0.00203<br>(0.00036) | $2.47 \times 10^{-8}$ | 0.321 | -0.00909<br>(0.00223) | $2.32 \times 10^{-5}$ | $4.63 \times 10^{-5}$ |
| **PDILT** | rs77924615 | 16:20,381,010 | G/A | 0.800 | -0.00980<br>(0.00039) | $4.38 \times 10^{-139}$ | 0.800 | -0.00958<br>(0.00044) | $1.52 \times 10^{-104}$ | 0.826 | -0.01030<br>(0.00281) | $1.27 \times 10^{-4}$ | $2.53 \times 10^{-4}$ |

<sup>1</sup>Trans-ethnic and European ancestry CKDGen results after subtracting the effect of 4661 CHRIS participants included in the CKDGen meta-analysis.
Abbreviations: Chr, chromosome; Pos, position; EA, effect allele; OA, other allele; EAF, effect allele frequency; b, coefficient of association; SE, standard error of b.
\*Replication of a variant in strong LD with the CKDGen lead variant: in these cases, the replicated and not the CKDGen lead variant is reported. See Supplementary Table S2 for lead variant comparisons.

### Mediation analysis

To identify possible reasons for observed larger effects, we conducted mediation analysis of the association between eGFRcrea and each of the 163 variants at the 11 loci across 70 quantitative health traits available in CHRIS (Supplementary Table S1). Adjusting the eGFRcrea-SNP association for each trait in turn (step 2 of the mediation analysis; see Methods) resulted in a substantial change of the eGFRcrea-SNP association coefficient in between 4 and 12 variant-trait associations per locus (Fig. 4). As expected, adjustment for the positive control SCr resulted in the nearly complete knockdown of the SNP effect at all loci (effect sizes toward null). Often, the effect change happened when adjusting for traits closely related to kidney function such as urate (*GAB2*, *IGF1R*, *PIP5K1B*, *SHROOM3*, and *STC1*), urinary creatinine (*SHROOM3*), and urinary albumin (*IGF1R*). Serum electrolytes such as magnesium (*SHROOM3*), corrected calcium (*PIP5K1B*), and sodium (*DDX1*, *GAB2*, *RSBN1L*, *SLC34A1*, and *STC1*) were also widely involved. Of note was the widespread effect of FT3, FT4, and the activated partial thromboplastin time (aPTT) across all loci. When adjusting for FT3 and FT4, the SNP effect on ln(eGFRcrea) was often completely altered. This behavior across all loci may suggest the presence of either a measurement artifact or a general related condition affecting the underlying population.

**Figure 4.**
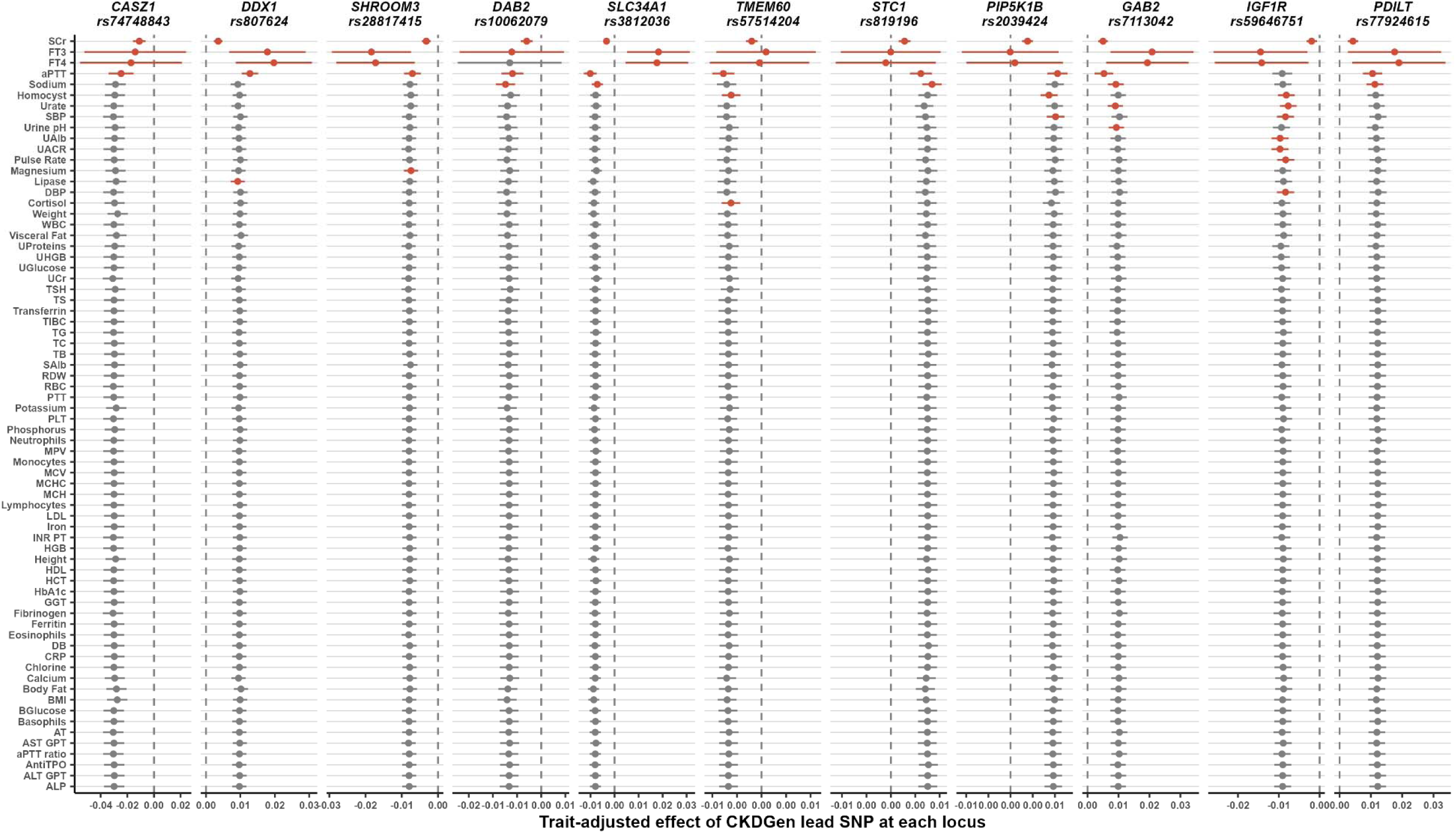
Results of the 2^nd^ step of the mediation analysis for the most associated variants at each locus. SNP effect size and their 95% confidence intervals are reported. Effects are expressed in terms of change of the natural logarithm of eGFRcrea in ml/min/1.73m^2^ per copy of the effect allele. Large departures from the expected effect size (‘outlier’, see Methods) are indicated in red. The vertical dashed lines identify the null. Large effect size changes were observed when adjusting for SCr (positive control; all loci), FT3 (all loci) and FT4 (10 loci), aPTT (10 loci), sodium (5 loci), homocysteine (3 loci), SBP (2 loci), urate (2 loci), and cortisol, DBP, lipase, magnesium, pulse rate, UACR, urinary albumin and urine pH (1 locus each).

In the 3^rd^ mediation analysis step, we evaluated whether the traits identified as effect modifiers in step 2 were associated with the SNP of which they modified the association with eGFRcrea (Table 3; Supplementary Table S4): SNP rs3812036 at *SLC34A1* was associated with aPTT levels (*P*=3.54×10^-6^) and five SNPs at *SHROOM3* were associated with serum magnesium levels (*P*-values between 5.33×10^-5^ and 6.12×10^-5^). Three of our associations reproduce previously reported associations found using Phenoscanner: at *SLC34A1*, Tang *et al.*^36^ reported association of rs3812036 with aPTT (*P*=2.88×10^-18^), and at *SHROOM3*, Meyer *et al.*^37^ reported associations of magnesium with rs4859682 (*P*=2.39×10^-9^) and rs13146355 (*P*=6.27×10^-13^).

**Table 3.**
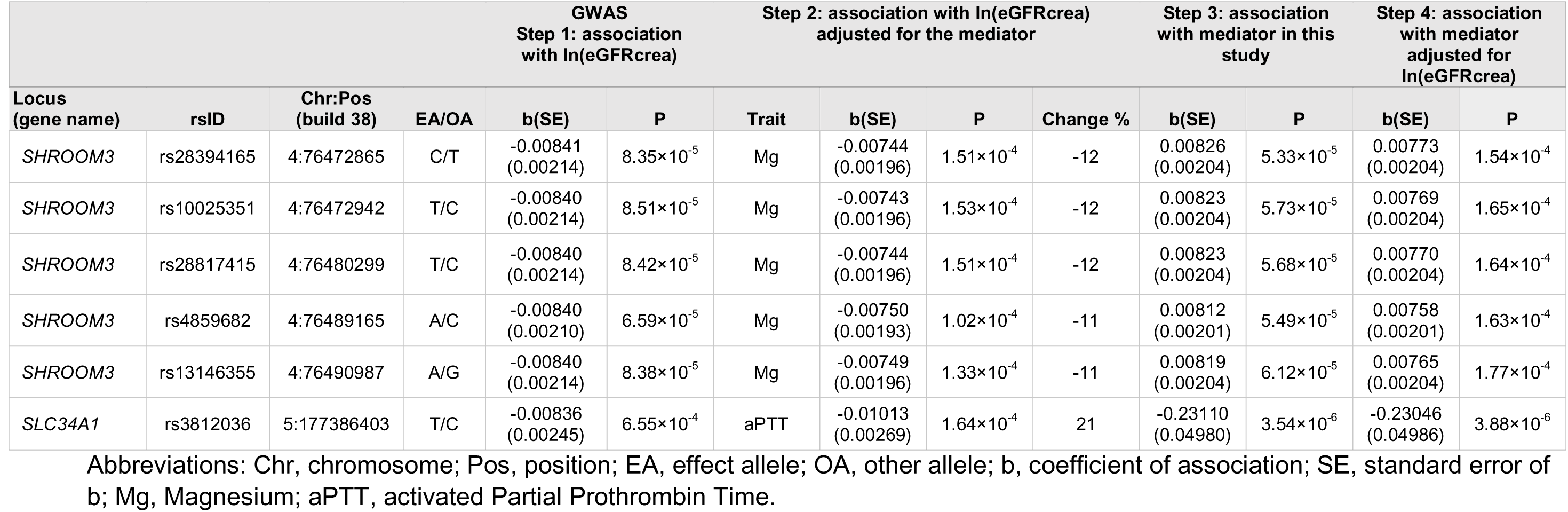
Results of the mediation analysis at *SHROOM3* and *SLC34A1*. Extensive mediation analysis results for all loci and all traits are reported in Supplementary Table S4.

All SNP-trait associations that successfully passed the 2^nd^ mediation analysis step were also interrogated against the Phenoscanner, GWAS Catalog, and ThyroidOmics consortium results (Methods) to identify associations that might have been missed by the 3^rd^ step of the mediation analysis in CHRIS due to lack of power (Supplementary Table S4). This analysis provided additional support for our findings at *SHROOM3* and *SLC34A1*. It also showed associations between variants at *IGF1R* and serum urate, unraveling how this additional mediation was likely missed by our study due to lack of power.

In summary, aPTT and serum magnesium qualified as partial mediators of eGFRcrea associations at *SLC34A1* and *SHROOM3*, respectively. At *SLC34A1*, the partial mediation of aPTT corresponded to a 21% larger effect of the SNP on ln(eGFRcrea); at *SHROOM3*, the partial mediation of serum magnesium corresponded to an effect attenuation of about 11% (Table 3).

### Interaction with thyroid dysfunction

Despite the pervasive effect change caused by FT3 and FT4 in step-2 analysis, these two traits did not result in significant step-3 associations and so did not qualify as mediators. However, motivated by the fact that hypothyroidism is amongst the leading causes of hospitalization in the region^38^, thyroid problems are common in Alpine areas, and the FT3 and FT4 adjustment affected the SNP-eGFRcrea association at all 11 loci, we considered it reasonable to evaluate a potential role of thyroid dysfunction as a modifier of the SNP-eGFRcrea relation.

Preliminarily, given that in the CHRIS study FT3 and FT4 were measured only in individuals with TSH levels above or below specific thresholds, we first conducted sensitivity analyses to exclude the presence of artifacts. We did not observe any genotype stratification by measured versus unmeasured FT3 and FT4 levels (Supplementary Table S5). Adjustment for municipality of residence, to exclude the presence of local clusters of thyroid dysfunction, did not alter the results (Supplementary Fig. 3).

We then tested for the presence of linear interaction with TSH levels, which identified a significant interaction at *STC1* SNPs rs819185 (*P*=0.00177) and rs819196 (*P*=0.00154) both below the multiple testing threshold of 0.05/11 loci (Fig. 5; Supplementary Table S6). Since for most loci the adjustment both for FT3 and for FT4 were causing an increase of the effect magnitude, we also postulated a U-shaped interaction model, with the SNP effects on ln(eGFRcrea) being larger both in hyper-and in hypothyroidism. We classified individuals as healthy (88.5%), with hypothyroidism (9.8%) or with hyperthyroidism (1.7%; Table 1). Our results did not support the presence of interaction with these two conditions at any locus after multiple testing control, despite nominally significant *P*-values observed for interaction between hyperthyroidism and SNPs in *SHROOM3* (*P*=0.03) and between hypothyroidism and SNPs in *PIP5K1B* (*P*=0.03) and *GAB2* (*P*=0.04; Supplementary Table S7).

**Figure 5.**
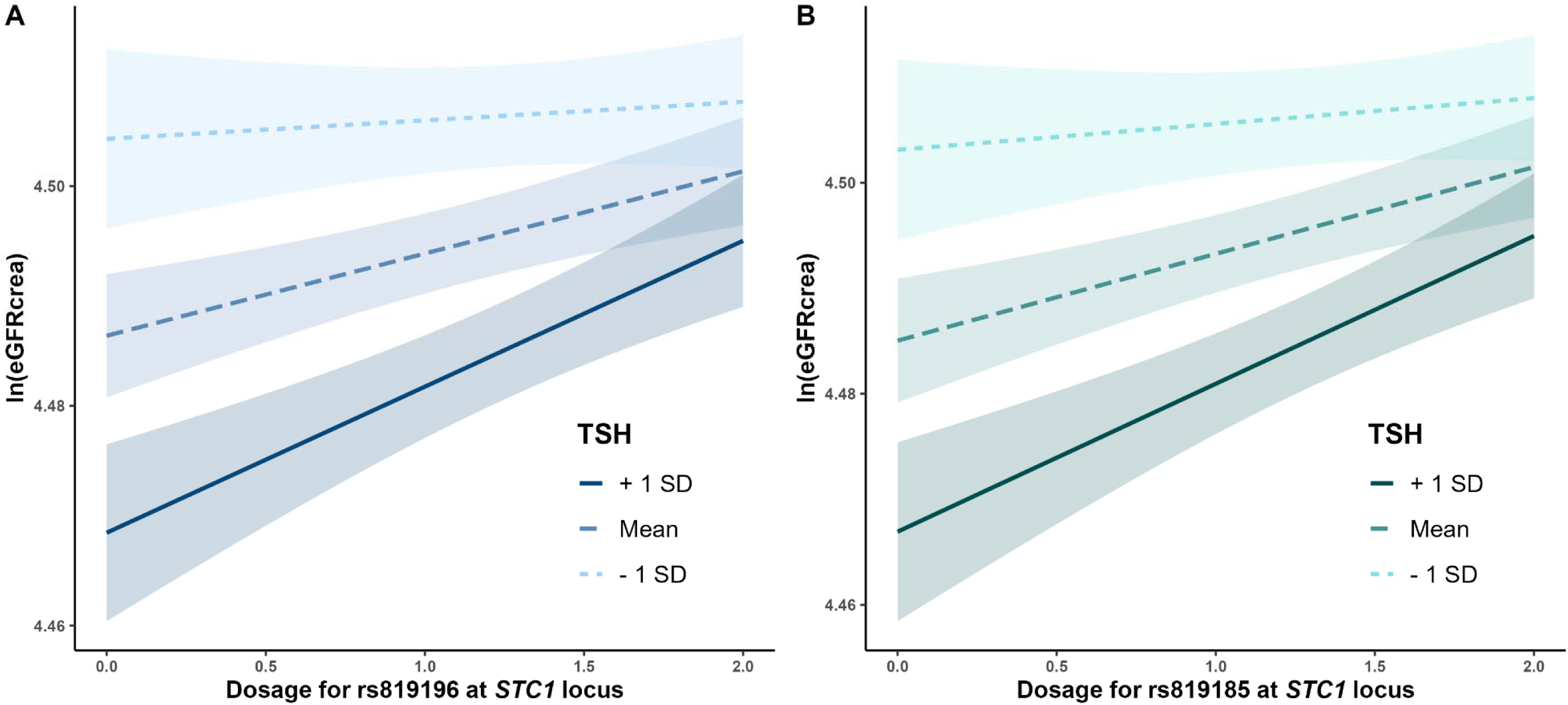
Interaction of ln(eGFRcrea)-associated variants with TSH levels. Effects of *STC1* SNPs rs819185 (P=0.00177; **Panel A**) and rs819196 (P=0.00154; **Panel B**) are larger at lower TSH levels.

## Discussion

Out of 147 loci known for their association with kidney function, our analysis identified 11 that were more strongly associated with eGFRcrea in the CHRIS study sample than in average European-ancestry population samples. Observed differences in effect magnitude could not be explained by MAF differences. Extensive mediation analysis identified serum magnesium and aPTT as partial mediators of the association of eGFRcrea with variants at *SHROOM3* and *SLC34A1*, respectively. SNP-by-TSH interaction analysis highlighted that the effect of variants at *STC1* on eGFRcrea would vary by TSH levels.

*PDILT*, *SHROOM3*, and *SLC34A1* explained the same fraction of eGFRcrea variance in CHRIS and CKDGen. In fact, for these loci, a relatively small sample size proved to be sufficient for their detection.^34,35^ For all other replicated loci, the replication was due to an interplay between allele frequency differences and effect estimate differences. Figures 3A and 3B show that the effect difference was probably more relevant than the allele frequency differences. But the question remains as to why precisely these and no other loci showed such large effects in CHRIS. SNPs may show larger effects at lower MAF. This situation was observed at just *DDX1* and *CASZ1*, whose effects got larger as the CHRIS-to-CKDGen MAF ratio got smaller. However, the MAF difference was unrelated to the larger effects observed at *GAB2*, *TMEM60*, *PIP5K1B*, and *IGF1R*: in these cases, MAF was similar or even smaller in CKDGen than in CHRIS (Fig. 2). The presence of un-modeled sample structure causing genomic inflation can be excluded as relatedness was appropriately modeled. We can also exclude the presence of genetic admixture as the CHRIS study was conducted in a small area, on a local population of homogeneous ancestry^15,39–41^. Differences in LD structure between the local and the overall population can also be excluded as regional association plots showed similar shapes between CHRIS and CKDGen. Epistatic gene-gene interaction would be a further possibility that cannot be easily verified. The most plausible remaining reasons would be the presence of local effect modifiers or gene-by-environment interaction.

Mediation analysis based on commonly measured biochemical and anthropometric traits provided limited answers in this respect but highlighted relevant mechanisms at three loci. The first finding was the identified presence of partial mediation of serum magnesium levels on the association of eGFRcrea with the *SHROOM3* locus. Genetic variants at *SHROOM3* have been associated with CKD^8,9^, reduced eGFRcrea^9^, increased albumin-to-creatinine ratio^10^, and low serum magnesium levels^11^. *SHROOM3* is necessary to maintain the glomerular filtration barrier integrity^42^. Variants in this gene have been shown to be associated with increased risk of CKD^43^, likely through disruption of the transcription factor TCF7L2 in podocyte cells. At the same locus, *FAM47E* is highly expressed in the human glomeruli and its transcription levels were associated with eGFRcrea^44^. Additional genes in this locus include *STBD1* and *CCDC158*, for which no connection with kidney function was demonstrated. The kidney is one of the primary regulators of serum magnesium concentration. It has been suggested that *SHROOM3* might increase serum magnesium levels through decreased renal magnesium excretion^45^. At the *SHROOM3* rs13146355 SNP, a previous GWAS on serum magnesium showed that the allele related with lower magnesium levels was associated with higher eGFRcrea levels^37^. The authors observed that the rs13146355 association with magnesium did not change when adjusting for eGFRcrea, suggesting pleiotropic independent effects. In our analysis, the effect of rs13146355 on eGFRcrea was reduced by 11% when adjusting for magnesium, suggesting partial mediation and thus the presence of partially overlapping mechanisms. Deeper analyses at the molecular level are warranted to identify the relevant mechanism.

A similar situation was observed at *SLC34A1* which, in addition to eGFRcrea^35^, was also previously associated with aPTT in a GWAMA of 9240 European ancestry individuals^36^ including participants from the same areas where the CHRIS study was sampled. By quantifying the clotting time from the activation of factor XII (*F12*) until the formation of a fibrin clot, aPTT is a measure of the integrity of the intrinsic and common coagulation pathways^46^. The observed mediation, in which adjustment for aPTT increases the SNP-eGFRcrea association by 21%, is likely related to the proximity between *SLC34A1* and the coagulation factor gene *F12*. The LD at this locus is compatible with the presence of common haplotypes tagging both genes, inducing pleiotropic independent effects on eGFRcrea and aPTT.

An additional relevant finding was the presence of an interaction between two SNPs at *STC1* and TSH levels. *STC1* is chiefly expressed in the thyroid gland (https://gtexportal.org/home/gene/STC1). It is also highly expressed in the kidney, where it is exclusively expressed in the collecting duct^47^. *STC1* is also associated with urate and urea levels and is involved in the physiological response to dehydration and protein-rich diet^47^. The *STC1*-encoded protein, stanniocalcin-1, is involved in phosphate reabsorption in the kidney proximal tubules and in multiple pathophysiological mechanisms including ischemic kidney injury^48^. Stanniocalcin-1 behaves like an endocrine hormone, with both autocrine and paracrine functions. Our observation of a SNP effect on eGFRcrea depending on TSH levels would suggest that altered thyroid function levels might affect Stanniocalcin-1 levels with consequent effect on kidney function phenotypes. While intriguing, this hypothesis should be verified by appropriate studies.

Our study had both strengths and limitations. The exploitation of a local homogeneous population sample was at the same time the most relevant strength and limitation of our study. While homogeneity may enhance the identification of specific genetic associations, it may also hinder the possibility to explain the underlying reasons of the observed larger effects compared to the average European ancestry population. The availability of a very large number of clinical parameters has allowed us to conduct an exhaustive and unbiased screening on potential mediation mechanisms. However, the analyzed traits lacked molecular specificity to identify underlying mechanisms. Given eGFRcrea might also reflect creatinine metabolism in addition to kidney function, another limitation of our study was the lack of additional kidney function markers such as cystatin C or BUN. For this reason, we limited our analyses to loci whose kidney function relevance was already demonstrated by previous studies^17^. Assessment of environmental exposures may also be a tool to explain the observed larger effects and would be ground for further research. Finally, our choice of adjustment for multiple testing is also worth a brief reflection. We have considered 147 loci that were already validated by at least one previous study through discovery and independent replication^17^, with several of the same loci being already reported previously^35,49,50^. As such, they wouldn’t need further validation and a nominal significance level would be commonly accepted for statistical testing. However, the aim of our analysis was to identify loci with substantially large effect in the CHRIS study to further conduct meaningful mediation analyses. For this reason, we choose to penalize the significance level by the number of independent loci. To us, this seems to be a sensible choice also by the fact that 147 independent tests covered 98% of the total variability of the 6337 SNPs in strong LD observed overall.

In conclusion, our study has identified loci that have particularly large effect on kidney function in the specific Alpine population sample considered in this study, namely *CASZ1*, *IGF1R*, *GAB2*, and *DDX1*. Allele frequency and LD analyses did not explain the observed larger effects nor did the extensive mediation analyses conducted. On the other hand, we showed that the effects of *SHROOM3* and *SLC34A1* on eGFRcrea are partially mediated by serum magnesium and aPTT, respectively. Finally, the observed *STC1*-TSH interaction implicates thyroid function involvement, which might have been favored by the high burden of thyroid-related diseases in the region. Further investigations are warranted to link the identified loci to environmental and molecular characteristics of specific population samples that may elucidate mechanisms not otherwise identifiable in large genetic meta-analyses.

## Supporting information

Supplementary tables revised

## Acknowledgements

The CHRIS study is a collaborative effort between the Eurac Research Institute for Biomedicine and the Healthcare System of the Autonomous Province of Bozen/Bolzano. We thank all study participants, the general practitioners, the personnel of the Hospital of Schlanders/Silandro, the field study team and the personnel of the CHRIS Biobank (BRIF code BRIF6107) for their support and collaboration. Extensive acknowledgement is reported at https://translationalmedicine.biomedcentral.com/articles/10.1186/s12967-015-0704-9.

## Author contributions

Conceptualization of the research project: C.P., D.G.

Recruitment and study management: M.G., P.P.P., C.P.

Bioinformatics: D.G., E.K., D.E., C.F.

Data quality control and harmonization: M.G., L.B., L.F.

Statistical Analysis: D.G.

Results interpretation: D.G., C.P., C.F., D.E., M.G., L.F., R.F.

Manuscript drafting: D.G., C.P.

Manuscript critical revision: D.E., E.K., R.F., L.F., L.B., M.G., D.J.M.P., P.P.P., C.F., D.G., C.P.

## Data availability

The data used in the current analyses can be requested with an application to at the Eurac Research Institute for Biomedicine.

## Supplementary Material

### Table of Contents

Supplementary method. Quantile normalization of the 70 health traits used in the mediation analysis 29

Supplementary Figure S1. Cumulative variance of 6337 variants over 147 loci explained by principal components in the CHRIS study. Adjusting for 147 independent tests covers 98% of the cumulative variance (red dot) 30

Supplementary Figure S2. Fitting PC-adjusted simple linear models came for practical reasons after observing substantial equivalence with kinship-adjusted linear mixed models obtained with EMMAX 31

Supplementary Figure S3. Quantile-quantile (QQ) plot of the P-values from the GWAS of ln(eGFRcrea) in the CHRIS study 32

Supplementary Figure S4. Regional association plots for the 11 replicated loci. Highlighted in purple is the most associated SNP in the CKDGen GWAS meta-analysis. All SNP positions are referred to the NCBI Build 38. Plots were generated with LocusZoom version 1.4 (33

Supplementary Figure S5. Comparison of effect size and standard error ratio for the variants in the model regressing ln(eGFRcrea) on age, sex, FT3 (or FT4), genetic variants with and without adjusting for municipality 39

**Supplementary method.** Quantile normalization of the 70 health traits used in the mediation analysis.

We used *normalize2Reference* function in the caret R package version 6.0-34 (the function is available in the later versions here: https://github.com/topepo/caret/tree/master/deprecated). Each trait was measured partially with an older method and partially with a newer, most recent method. The observations from the newer method (method = 1) were set as the reference to derive the quantiles of the trait distribution on which to standardize the observations from the older method (method = 0). We then replaced the observation from the older method with the standardized ones and included together with the values from the newer method.

### Notation

-**trait**: the quantitative variable to which quantile normalization was applied;
-**method:** the newer method for measuring the trait (1); the older method (0);
-**df:** dataframe; the data in which the trait of interest is available

library(caret)

quantile_norm <-normalize2Reference (data = df[df$method == 0, "trait"], refData = quantile(

df[df$method == 1, "trait"],

probs = seq(0, 1, length.out = length(df[df$method == 0, "trait"])),

na.rm = TRUE, names = TRUE, type = 7,

digits = 7), ties = TRUE)

\# Creating a new variable for the quantile normalized trait

df$trait_std <-df$trait

\# Replacing non-transformed values with the quantile-normalized values

df[df$method == 0, "trait_std"] <-quantile_norm

---

**Supplementary Figure S1.**
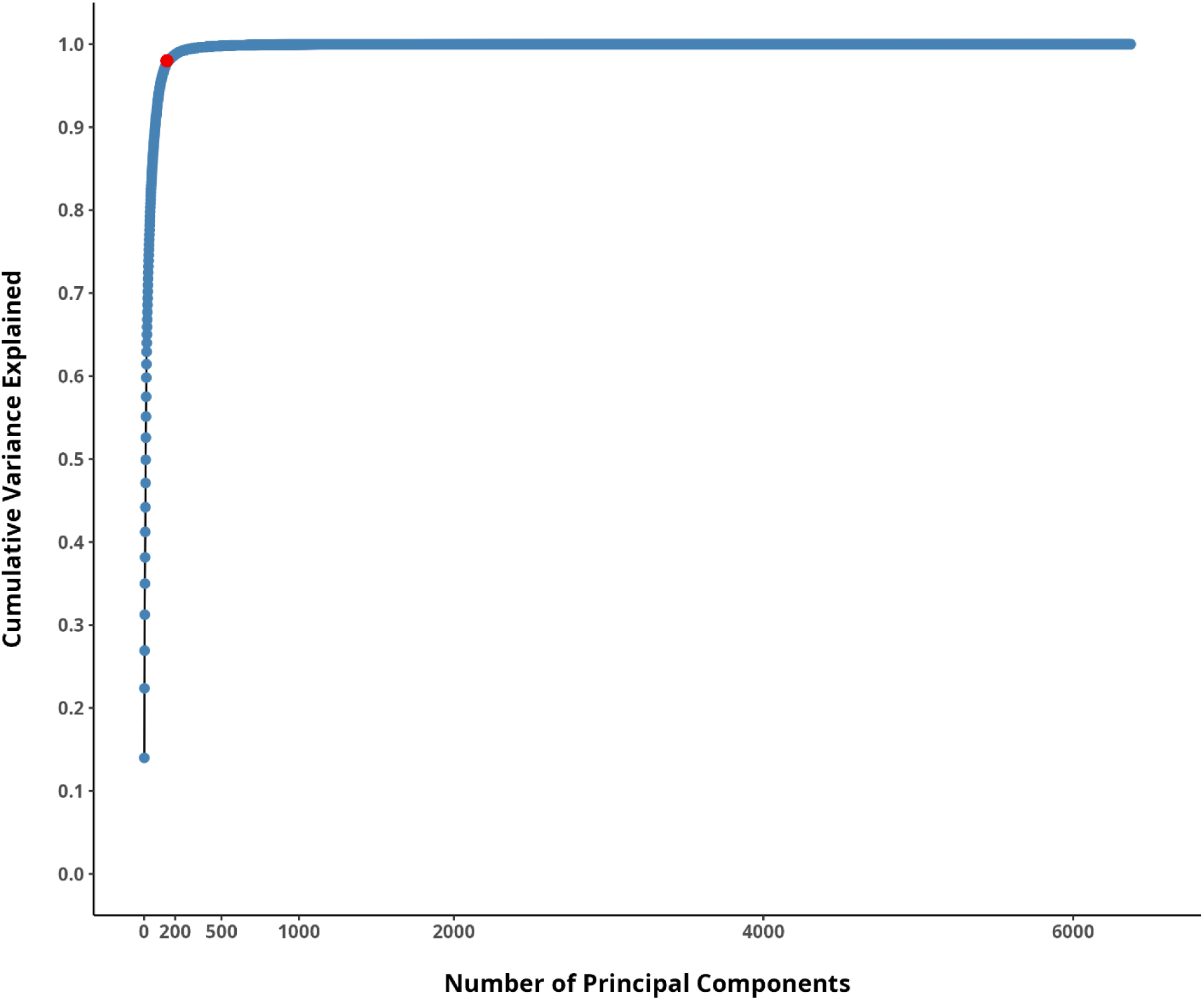
Cumulative variance of 6337 variants over 147 loci explained by principal components in the CHRIS study. Adjusting for 147 independent tests covers 98% of the cumulative variance (red dot).

**Supplementary Figure S2.**
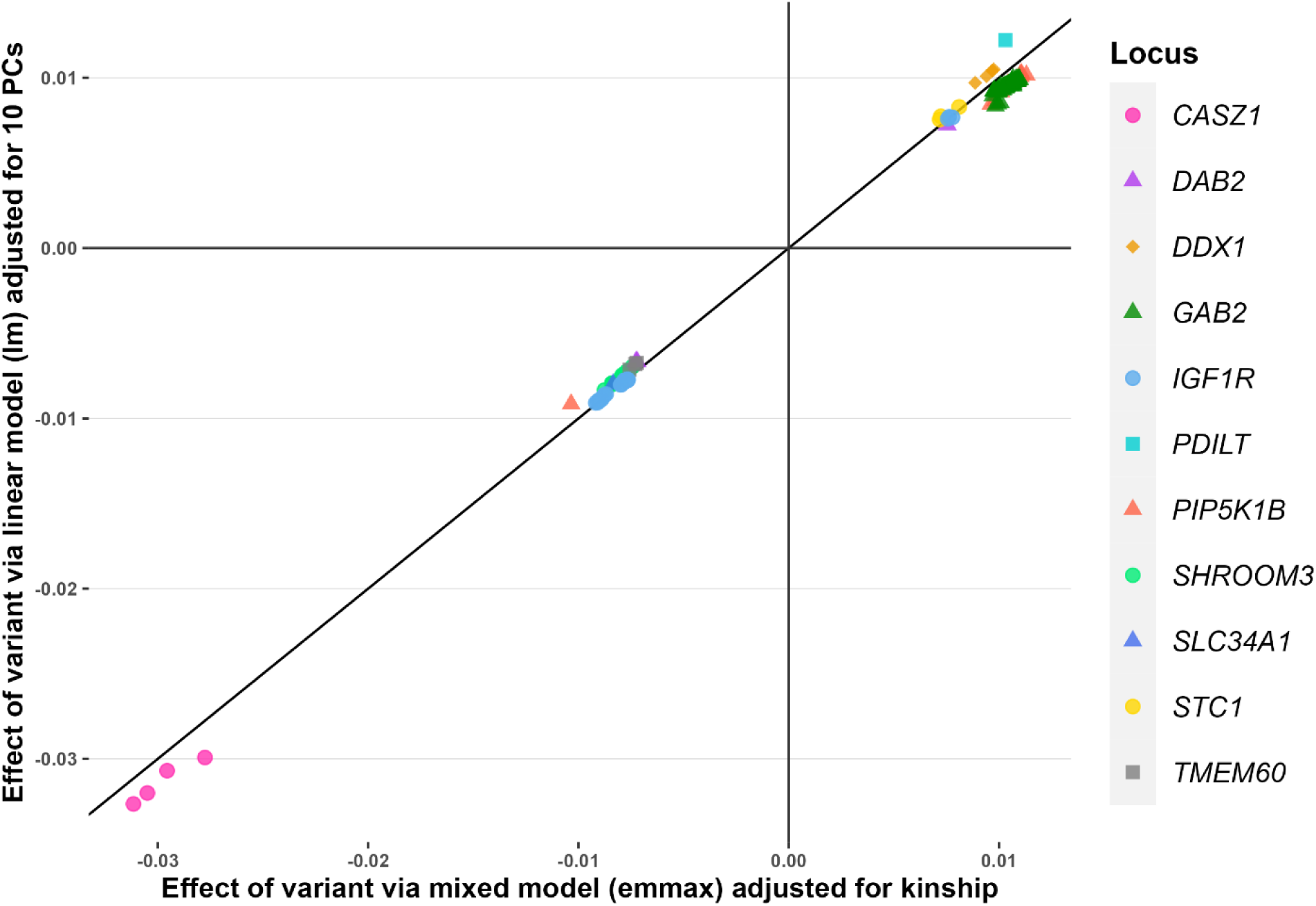
Fitting PC-adjusted simple linear models came for practical reasons after observing substantial equivalence with kinship-adjusted linear mixed models obtained with EMMAX.

**Supplementary Figure S3.**
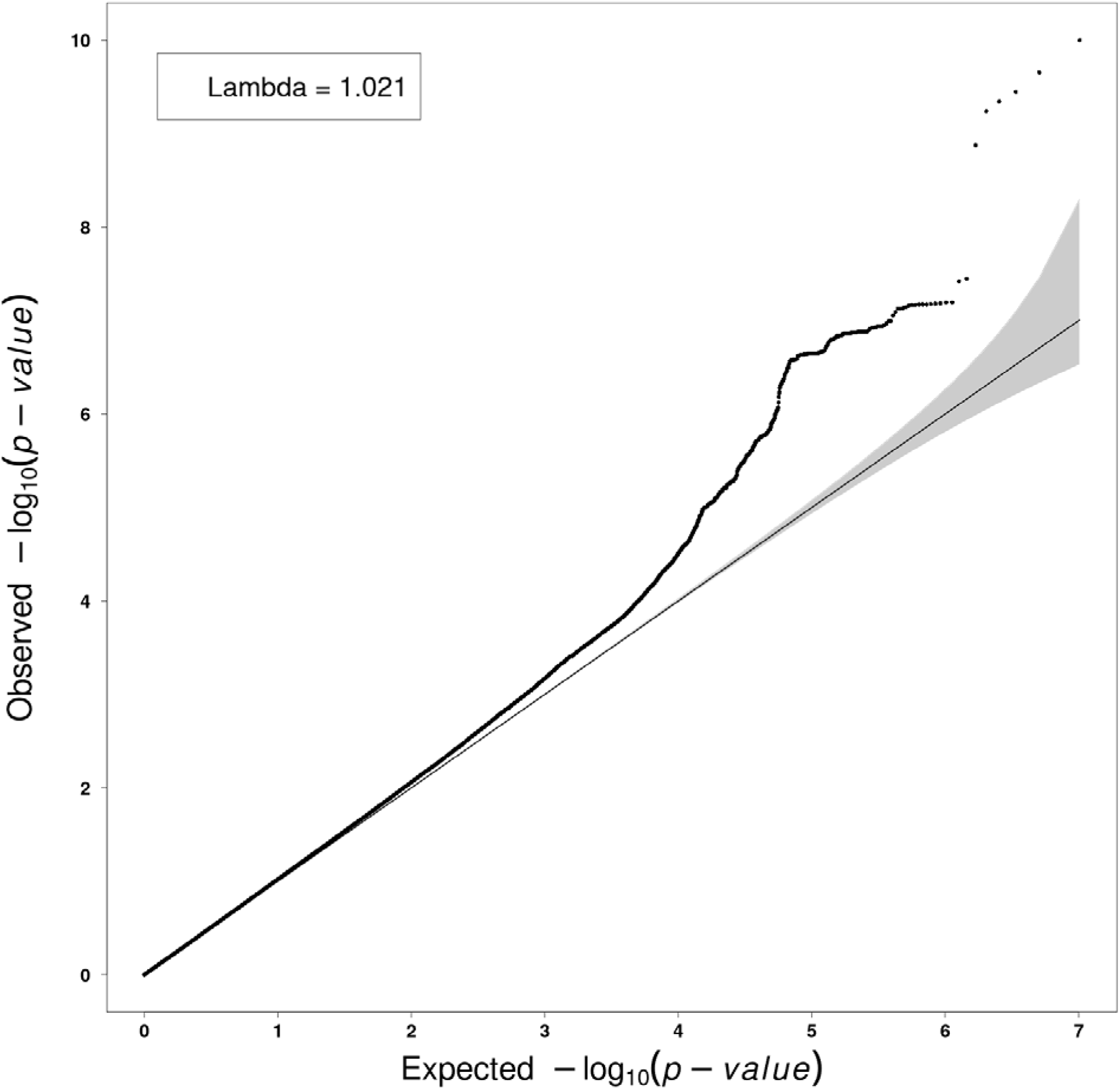
Quantile-quantile (QQ) plot of the P-values from the GWAS of ln(eGFRcrea) in the CHRIS study.

**Supplementary Figure S4.**
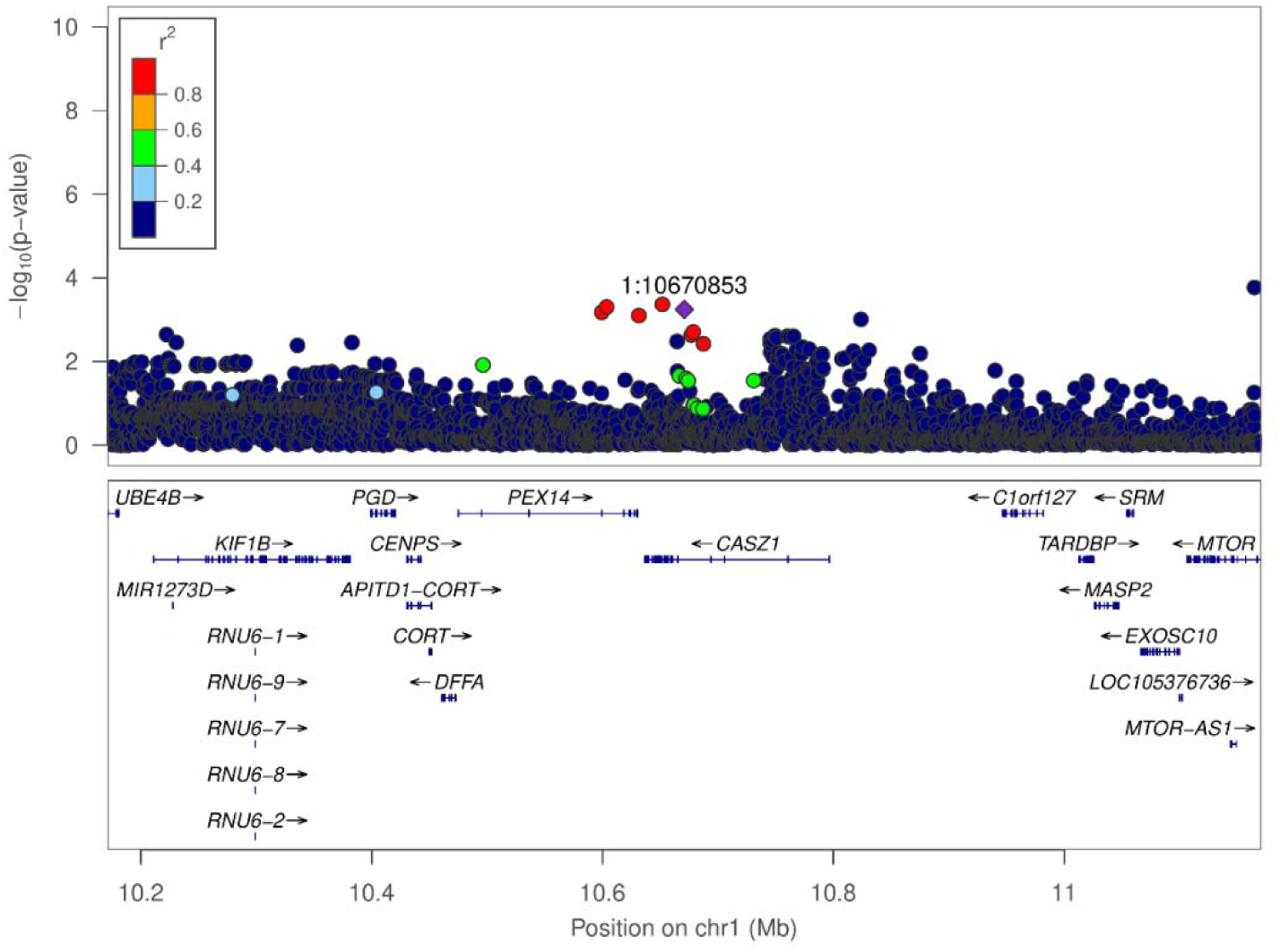
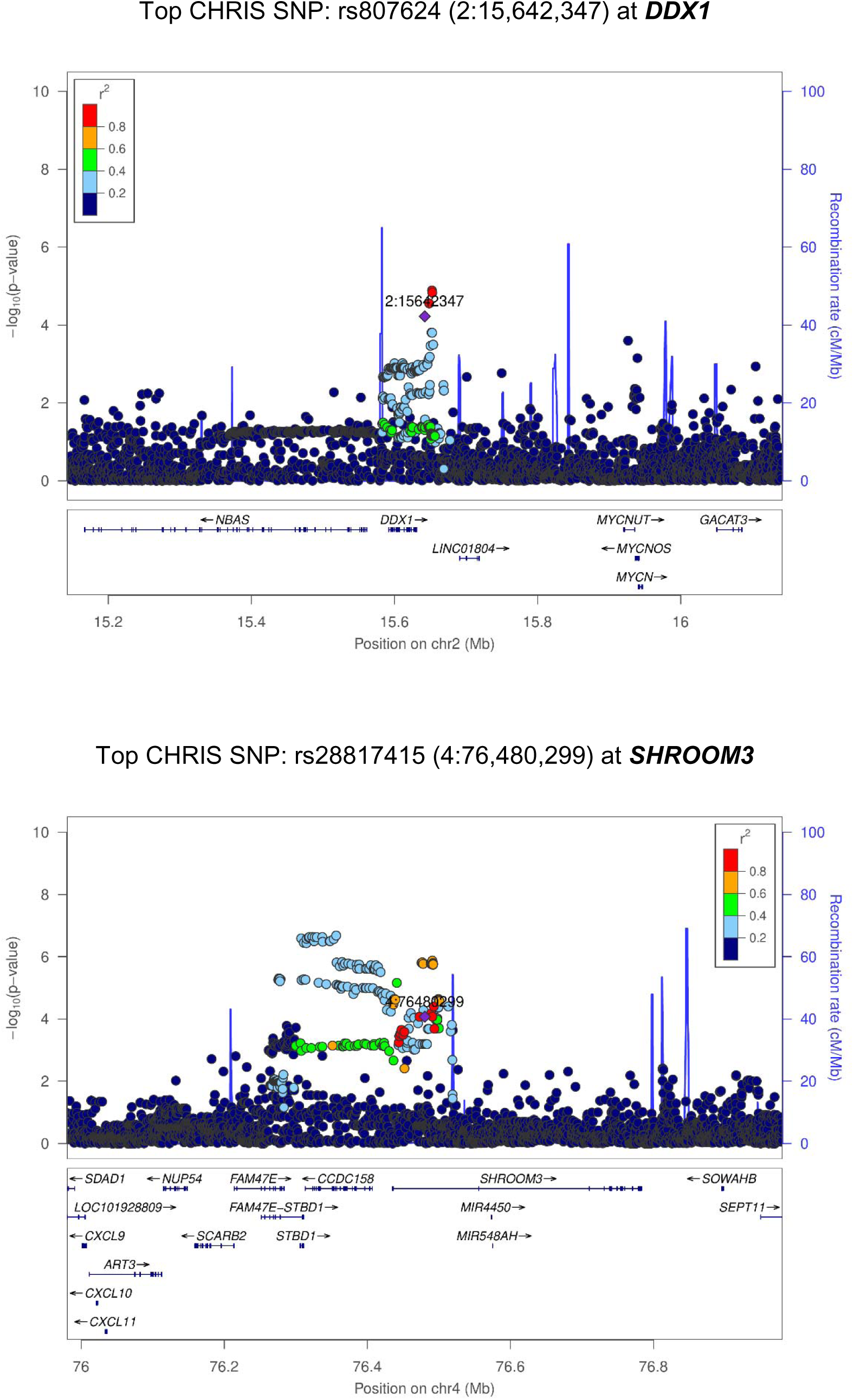
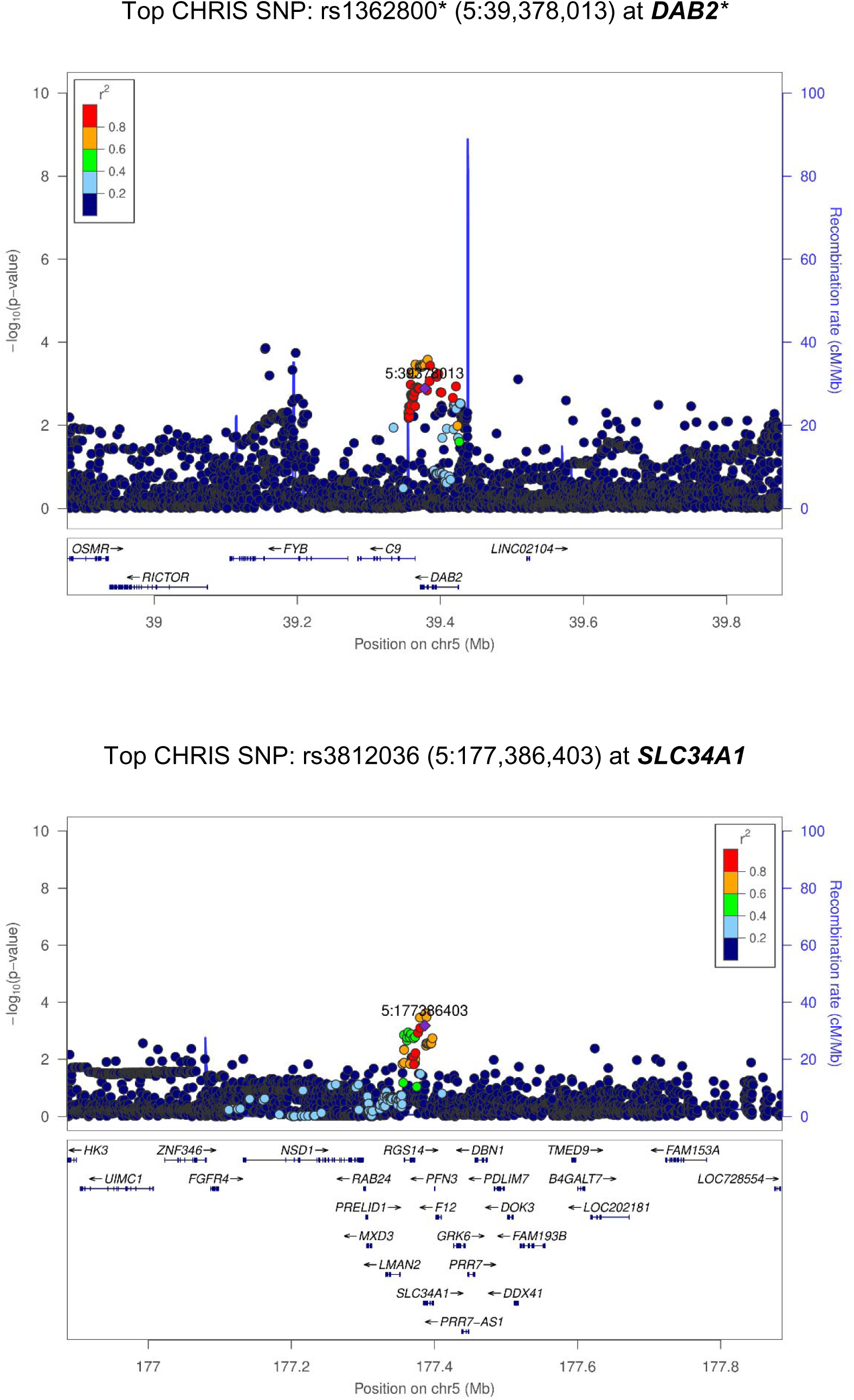
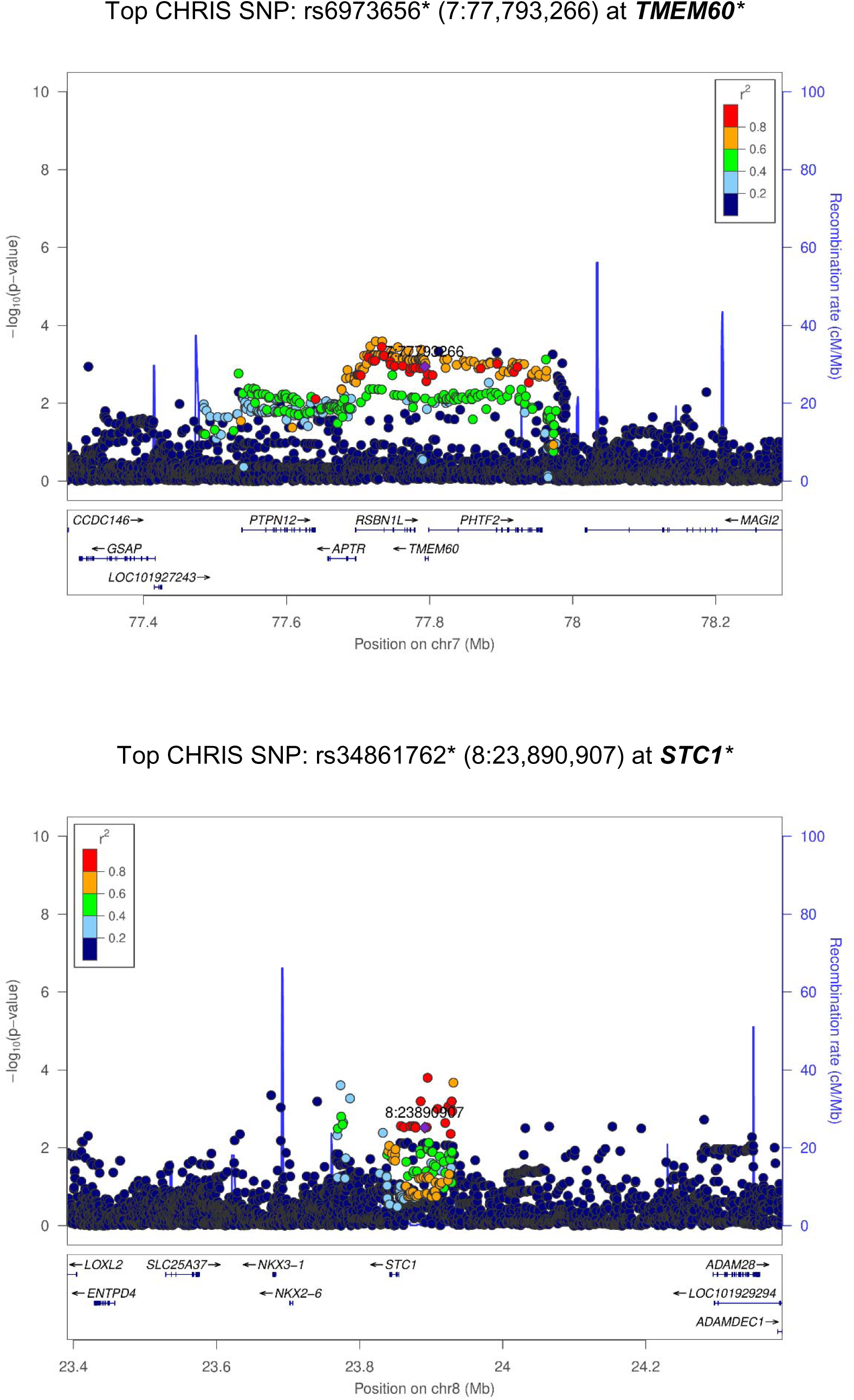
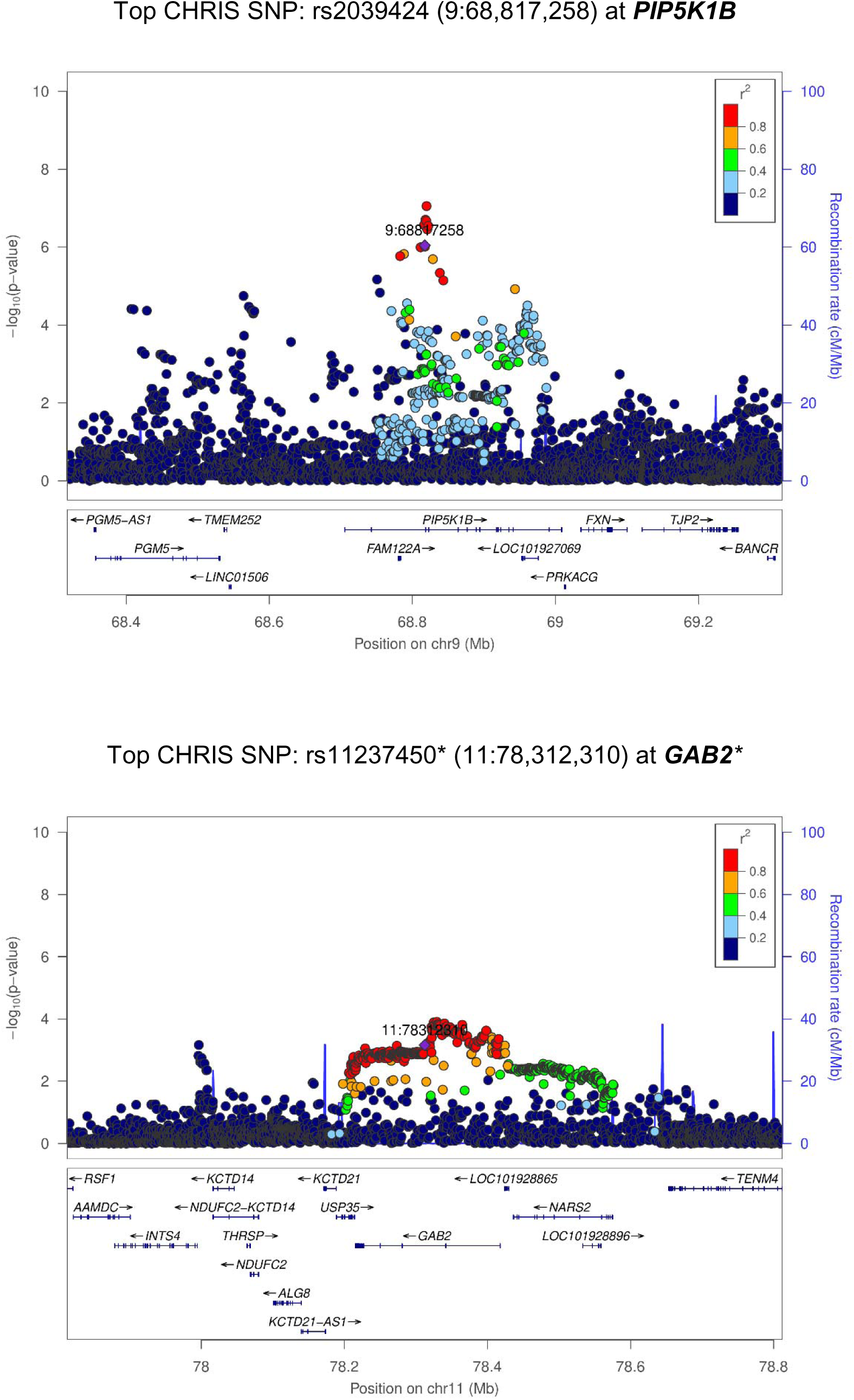
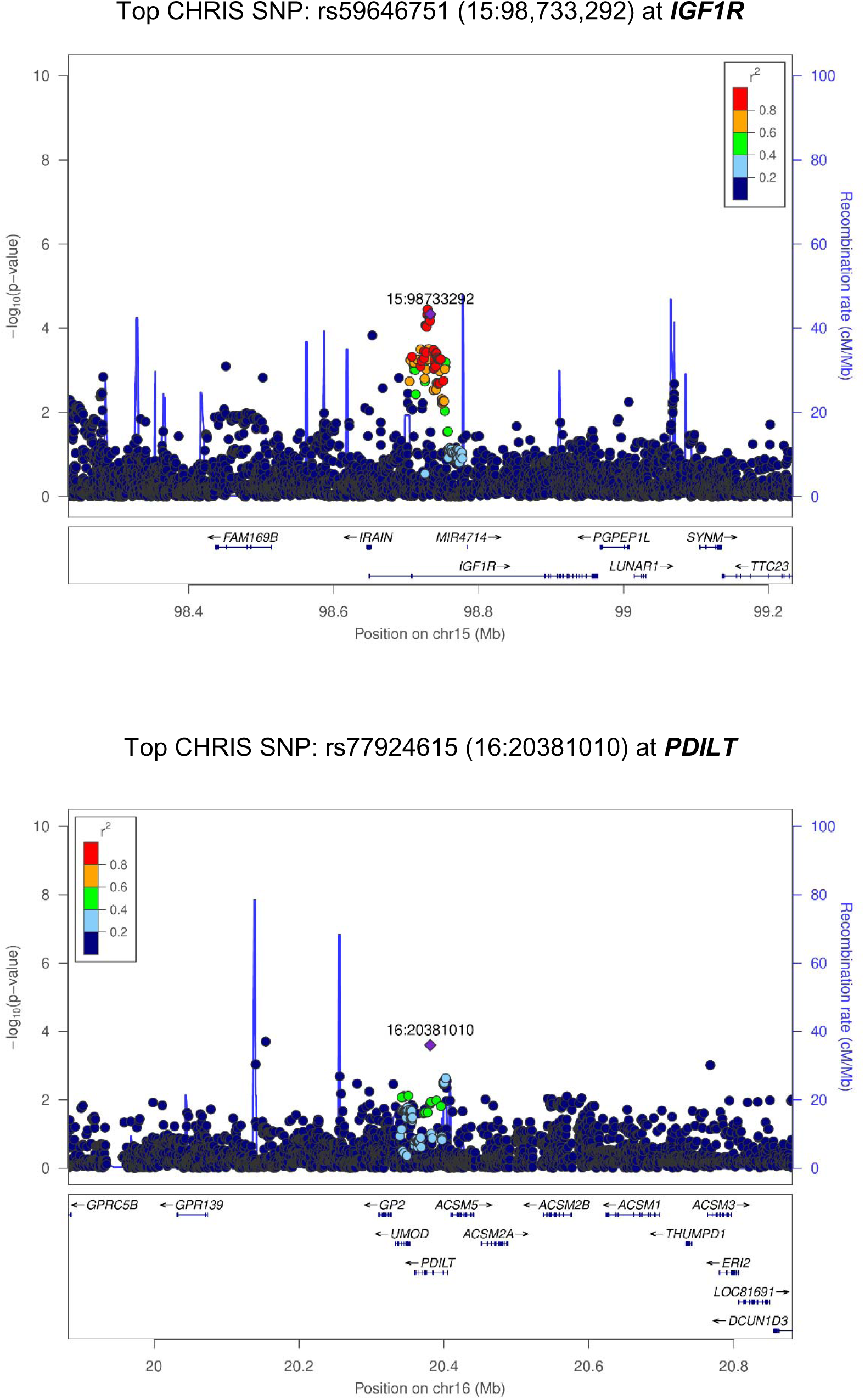
Regional association plots for the 11 replicated loci. Highlighted in purple is the most associated SNP in the CKDGen GWAS meta-analysis. All SNP positions are referred to the NCBI Build 38. Plots were generated with LocusZoom version 1.4 (Pruim RJ, et al., Bioinformatics, 2010. 26(18): 2336-7). * indicates cases where not the CKDGen lead SNP but a proxy of it was replicated in the CHRIS study.

**Supplementary Figure S5.**
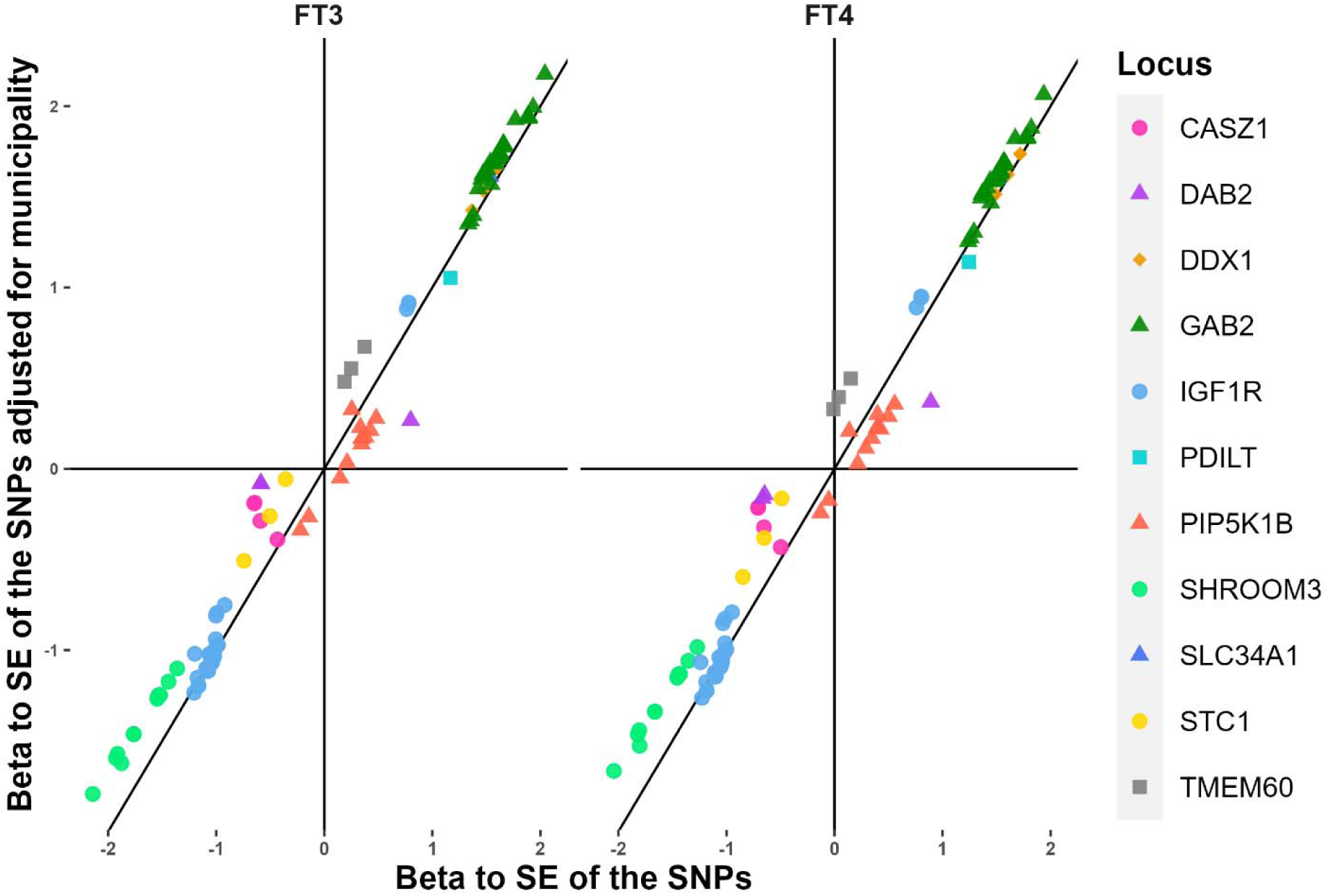
Comparison of effect size and standard error ratio for the variants in the model regressing ln(eGFRcrea) on age, sex, FT3 (or FT4), genetic variants with and without adjusting for municipality.

